# A “step too far” or “perfect sense”? A qualitative study of British adults’ views on mandating COVID-19 vaccination and vaccine passports

**DOI:** 10.1101/2022.02.07.22270458

**Authors:** Martine Stead, Allison Ford, Douglas Eadie, Hannah Biggs, Claire Elliott, Michael Ussher, Helen Bedford, Kathryn Angus, Kate Hunt, Anne Marie MacKintosh, Curtis Jessop, Andy MacGregor

## Abstract

**Background:** Debate is ongoing about mandating COVID-19 vaccination to maximise uptake. Policymakers must consider whether to mandate, for how long, and in which contexts, taking into account not only legal and ethical questions but also public opinion. Implementing mandates among populations who oppose them could be counterproductive.

**Methods:** Qualitative telephone interviews (Feb-May 2021) with British adults explored views on vaccine passports and mandatory vaccination. Participants (n=50) were purposively selected from respondents to a probability-based national survey of attitudes to COVID-19 vaccination, to include those expressing vaccine-hesitancy. Data were analysed thematically.

**Findings:** Six themes were identified in participants’ narratives concerning mandates: (i) mandates are a necessary and proportionate response for some occupations to protect the vulnerable and facilitate the resumption of free movement; (ii) mandates undermine autonomy and choice; (iii) mandates represent an over-reach of state power; (iv) mandates could potentially create ‘vaccine apartheid’; (v) the importance of context and framing; and (vi) mandates present considerable feasibility challenges. Those refusing vaccination tended to argue strongly against mandates. However, those in favour of vaccination also expressed concerns about freedom of choice, state coercion and social divisiveness.

**Discussion:** To our knowledge, this is the first in-depth UK study of public views on COVID-19 vaccine mandates. It does not assess support for different mandates but explores emotions, principles and reasoning underpinning views. Our data suggest that debate around mandates can arouse strong concerns and could entrench scepticism. Policymakers should proceed with caution. While surveys can provide snapshots of opinion on mandates, views are complex and further consultation is needed regarding specific scenarios.

## Background

Vaccination is vital for managing COVID-19. Mandating vaccination to maximise uptake is contentious [1–3]. Policymakers must consider whether to mandate, for how long, and in which contexts, taking into account not only legal and ethical questions but also public opinion [4].

Governments sometimes mandate behaviours to reduce harm (e.g., wearing seatbelts, lockdowns). Mandatory vaccination could be ethically justified to protect vulnerable people, progress towards herd immunity, and spread the burden of reaching herd immunity [3,5,6]. However, mandates reduce liberty and autonomy, raising ethical concerns which need to be balanced with public health goals [1]. The World Health Organization has identified several ethical issues to be considered before mandating COVID-19 vaccination, including effects on public trust and confidence [1]. Implementing mandates among populations who oppose them, particularly the vaccine hesitant, could be counterproductive [4,7]. Mandates could increase vaccine hesitancy and resistance [8], and be ‘weaponised’ by anti-vaccination movements [5,7].

Few governments are considering mandatory vaccination of the general population. In Brazil local governments can make COVID-19 vaccination mandatory, although citizens cannot be physically forced to be vaccinated [9]. Mandatory vaccination in specific contexts is being widely considered, such as for health and social care workers in direct contact with people at high risk [1,2,8,10–12]. Italy made COVID-19 vaccination mandatory for healthcare workers on 1^st^ April 2021 [13]. Ireland contemplates mandatory vaccination of healthcare workers but considers it “intrusive” [14]. In England, COVID-19 vaccination became mandatory for care home staff in July 2021, but opinion is divided [15–17]. Consultations are planned on mandating COVID-19 and flu vaccinations for UK National Health Service staff [18].

Requiring proof of vaccination (e.g. ‘vaccine passports’ linking vaccination status to identity) for access to certain activities is a form of mandate [19,20], and may encourage uptake [21]. Vaccine passports have been discussed primarily for international travel, but also for education, workplaces, entertainment and hospitality [20,22–24]. Israel introduced short-term vaccine passports in March-June 2021 [21]; such schemes continue to be debated internationally. There are concerns about the potential effectiveness, practicality, security, legality, ethicality and acceptability of vaccine passport schemes and their potential to exacerbate inequalities [7,19,20,23].

Surveys suggest divided public opinion on vaccine mandates, with variation by mandate type and country [6,23,25–28]. If mandates are introduced, public acceptability is essential. To our knowledge, no qualitative studies have examined public attitudes to COVID-19 vaccine mandates, contexts in which mandates are more or less acceptable, and bases for concerns and objections. We conducted such a study, focussing on those hesitant about vaccination.

## Methods

### Sample and recruitment

Participants (n=50) were from the OPTIMUM survey of adult (aged 18+) attitudes to COVID-19 vaccination in Great Britain (GB) (January-February 2021). The survey was administered to the probability-based NatCen Panel [29], recruited from the 2018, 2019, and 2020 waves of the British Social Attitudes survey, with respondents randomly selected from England, Wales and Scotland. Among 5,931 panellists invited, 84% responded. Those agreeing to further interview provided two purposive sub-samples (OPTIMUM GB (n=30) and OPTIMUM Boost Scotland (n=20)) for one-to-one interviews.

For both sub-samples, participants were selected according to quota controls and their survey responses to intent to be vaccinated (‘yes’, ‘no’, ‘unsure’). Both sub-samples were purposively skewed towards those who indicated vaccine hesitancy in the survey: the GB sample included 23 indicating hesitancy (16 ‘unsure’, 7 ‘refuse’); all of the Scotland sample did so (16 ‘unsure’, 4 ‘refuse’). To ensure a spread of demographic factors, minimum/maximum quotas were placed on age, sex, ethnicity, education, and country of residence. Potential participants (n=136) were emailed an information sheet and consent form by NatCen and followed-up by email/telephone to confirm participation, record consent (email or verbally) and arrange an interview. The achieved sample comprised 22 males and 28 females aged 18-70+, evenly distributed across (Scottish) Index of Multiple Deprivation quintiles. Forty-three participants were of white ethnicity and 19 were educated to degree level. Twenty-four were resident in England, 24 in Scotland and 2 in Wales (see Tables 1 and 2). Although we purposively recruited a majority who indicated hesitancy in their survey responses, by the time of interview some participants had accepted, or said that they would likely accept, vaccination (Table 2).

**Table 1.** Sample

| **Age** | **n** |
| --- | --- |
| 18-29 | 5 |
| 30-49 | 19 |
| 50-69 | 17 |
| 70+ | 9 |
| **Sex** |  |
| Male | 22 |
| Female | 28 |
| **Ethnicity** |  |
| White | 43 |
| Black, Asian & Minority Ethnic | 7 |
| **Country of residence** |  |
| England | 24 |
| Scotland | 24 |
| Wales | 2 |
| **Highest educational qualification** |  |
| Degree or equivalent, and above | 19 |
| A levels or vocational level 3 or equivalent and above, but below degree | 13 |
| Other qualifications below A levels or vocational level 3 or equivalent | 6 |
| Other qualification | 5 |
| No qualifications | 7 |
| **(S)IMD quintile<sup>#</sup>** |  |
| 1 | 9 |
| 2 | 12 |
| 3 | 9 |
| 4 | 12 |
| 5 | 8 |
| **Vaccine intent at survey** |  |
| Accept | 7 |
| Refuse | 11 |
| Unsure | 32 |

<sup>#</sup> (S)IMD = (Scottish) Index of Multiple Deprivation, 1 = most deprived and 5 = least deprived

**Table 2.**
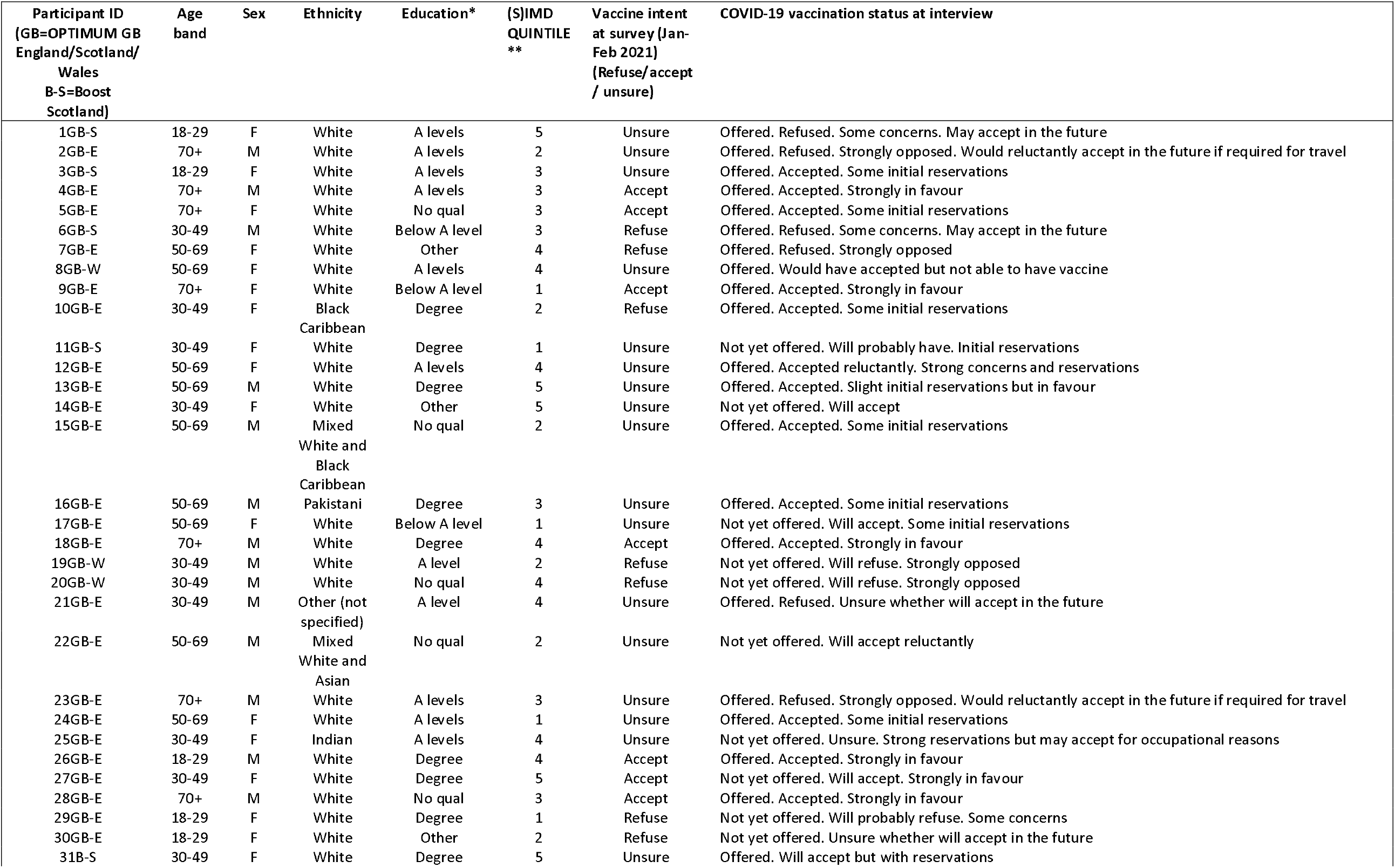
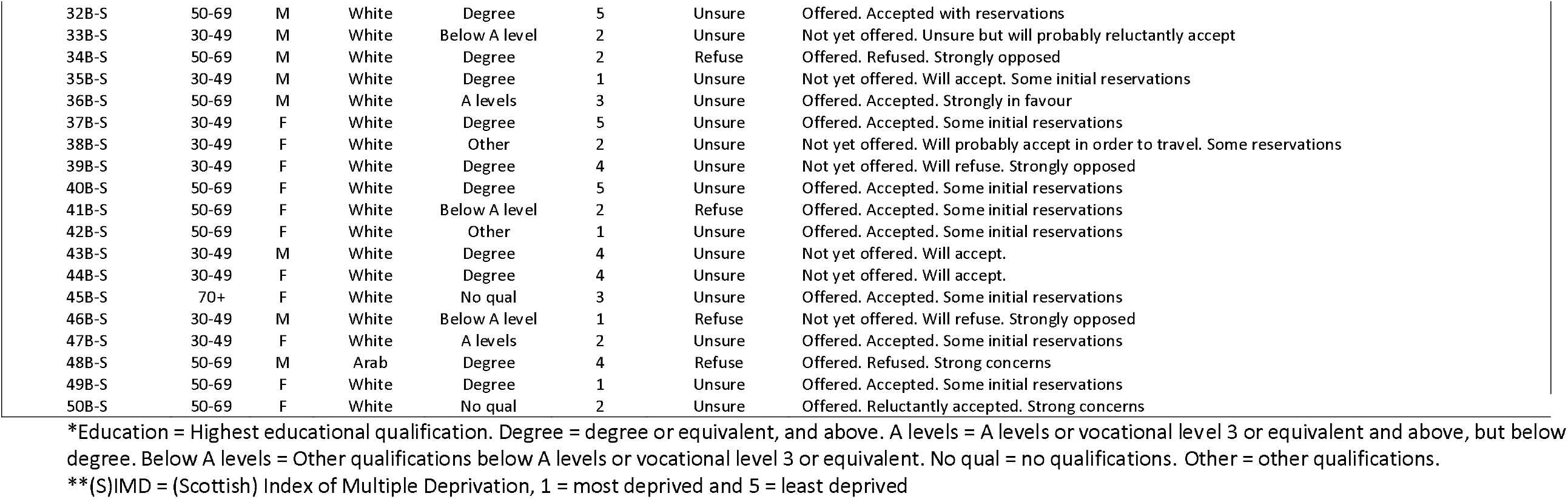
Individual participant characteristics

### Data collection

Semi-structured interviews (41-110 minutes) were conducted via telephone by MS, AF, DE, HB, IM, AM, and CE. The topic guide covered: background; COVID-19 beliefs; COVID-19 vaccine uptake expectations/intentions including motivations/concerns and other facilitators/barriers; knowledge and understanding of COVID-19 vaccine(s) and vaccination generally; information sources; social and community norms/expectations; and future possible mandates. For discussion on mandates, participants were asked for their views and feelings on passports and mandatory vaccination, advantages or disadvantages to mandates, and if vaccination should be compulsory for any groups. Some questions were amended to reflect the Scottish context. Participants received a £30 retail voucher (‘Love2Shop’) in recognition of their time. Interviews were conducted February-March 2021 for the GB sample and April-May 2021 for the Scotland sample. Interviews were audio-recorded and transcribed verbatim and anonymised.

### Analysis

Thematic analysis was deductive, informed by the topic guide, and inductive from participants’ accounts [30]. Key themes and issues were initially identified through familiarisation with transcripts. A coding framework was developed and refined by MS, AF, and DE who independently tested the framework on six transcripts. Using NVivo 12, transcripts were coded by AF, DE, HB, CE and AM. After coding, each coder prepared a summary of the transcript. MS, AF and DE subsequently met to agree each participant’s vaccination status and attitude (Table 2). For data coded under the mandates theme, an initial summary was prepared, including key quotes, followed by a detailed analysis, developing key themes underpinning participants’ attitudes. These themes were refined, interpreted and labelled through discussion with the wider team.

## Findings

As similar themes emerged for different mandates, findings are presented by six overarching themes, rather than by type of mandate: (i) necessary and proportionate response; (ii) autonomy and consent; (iii) overreach of state power; (iv) vaccine apartheid; (v) fluidity of support and (vi) concerns about feasibility.

### (i) Necessary and proportionate response

Narratives under this theme endorsed the idea that mandatory vaccination in some contexts was a necessary and proportionate response in controlling the pandemic and protecting others. Generally, participants who expressed these views were in favour of COVID-19 vaccination and had accepted, or intended to accept, the vaccine.

Support for <u>universal</u> mandatory vaccination was expressed by only a handful of participants: “*It should be compulsory for everyone, to eradicate it completely.” (42B-S); “I think the vaccination should be compulsory, to be honest, to [sic] everyone…my understanding of what vaccination is, if you take the vaccine it’s [to] protect you from the virus.” (22GB-E)*. However, the idea of <u>targeted</u> mandatory vaccination was more widely acceptable. It was suggested this could apply to those particularly vulnerable to COVID-19 – “*probably those with underlying health conditions…it would make perfect sense to, for own protection” (9GB-E)* - and also to occupational groups whose direct contact with the public put them at risk of contracting or transmitting the virus: *“people who work with people in nursing homes, nurses, doctors. I would say keep those also safe…maybe teachers” (43B-S); “people who travel a lot say, as part of their job or whatever*…*because it protects, not just themselves, but everybody that they come into contact with” (9GB-E); “all the emergency services and teachers. They’re out dealing with the public and dealing with, you know, everything every day, aren’t they?” (14GB-E)* Some with vulnerable family members expressed the desire to protect them, such as an elderly relative at potential risk from unvaccinated carers *(40B-S)*, or a relative working in an occupation where unvaccinated colleagues could expose them to infection *(17GB-E)*. Some narratives robustly endorsed the logic of ‘no jab, no job’ - if someone was not prepared to accept the vaccine then they could not work in a particular sector: *“I think if you’re going to work in the care sector like that and you refuse the injection because of, ‘I don’t want it and don’t believe it’ nonsense, then you shouldn’t be allowed…to be working with vulnerable people.” (47B-S) “My (relative) …some of the nurses there…would not have the injection…, as far as I’m concerned, they can infect my (relative)…If you don’t like it, then don’t have a job in a (care setting).” (28GB-E)*

Similarly, some saw vaccine certification as a condition of access to some activities and services as reasonable and proportionate. Firstly, there was perceived to be precedents for health and other forms of certification being required for travel: *“a lot of time, you need to prove that you have certain vaccination to enter the country, so I can’t see any problems with it” (31B-S)*; “*it’s the same if you went to Africa [e.g. for yellow fever]” (CV25); “I would never irk [sic] at showing an ID, I…I just feel, why not?*…*I just don’t understand why people need to get so upset about it.” (5GB-E)* Secondly, passports were justified if they speeded up the resumption of freedom of movement: *“It’s the easiest way to help the travel industry I think, because then having that passport means…we can all travel soon again” (44B-S); “As far as I’m concerned, if it means that I can go to see my (child) in (name of city) quicker,…from a selfish point of view, I’m cool about it.” (40B-S)* Thirdly, because holidays were a discretionary activity, the principle of choice was not undermined: *“I want to travel, I have to have a vaccine. It’s a choice.” (49B-S)* Following from this point, some argued that it was reasonable to withhold certain privileges from *“selfish” (42B-S)* people who chose not to share the societal burden of vaccination: *“If they are so stupid they’re not gonna have the vaccination, then don’t put other people at risk. So if you don’t want the vaccination, then don’t go on a flight, don’t go in the hospital to be treated, don’t go into a pub or restaurant because you’re gonna…you could infect other people.” (28GB-E)*. It was also suggested that a vaccine passport scheme could act as an incentive for vaccination: *“my natural reaction is that vaccine passports are actually a good idea…it’s rewarding good behaviour, rather than punishing bad behaviour, which I think is a lot better way to do things.” (26GB-E)* Support for this possibility was provided by some refusers: *“For me, that’s the only reason I would take it. If it meant to go abroad I needed a* [vaccine] *passport*…*But I don’t agree with it.” (38B-S)*

### (ii) Autonomy and consent

This theme positioned vaccine mandates, including universal mandatory vaccination, mandatory vaccination for particular occupations and vaccine passports, as a threat to autonomy, freedom of choice, and individual rights. These narratives sometimes used emotive language – *“disgusting” (23GB-E)* - indicating strength of discomfort and opposition: *“This passport that they’re thinking about, which I think is absolutely appalling, because you don’t get a passport say for cancer. You’re not going around with this big passport saying, ‘I’ve had cancer, this is all the treatments I’ve had’. Why do we want to do it for COVID?” (39B-S)*

Generally, participants who had refused or said they would refuse vaccination, or expressed strong reservations about it, were more likely to express such sentiments than those favouring vaccination. However, some who supported vaccination also held to the principle that it should never be compulsory. The primacy of choice was emphasised, even where the participant recognised potential conflict with their own support for vaccination: “*Oh…I’m not sure about that one…Oh dear. Because I feel that I approve of it and I’ve had it, I shouldn’t force my opinion onto other people.” (5GB-E)* In several narratives, including from those in favour of vaccination, freedom of choice and the principle that acceptance of any medical intervention should be based on informed consent trumped other considerations: “*no is the answer to that. I still think you’ve got to have, vaccination by consent” (13GB-E)*; “*You can’t make someone put a chemical into their body.” (4GB-E)* This included where it was recognised that vaccination could be *“in the best interests of the individual concerned” (21GB-E)* or would help to reduce the risk to people in their care if they worked in particular occupations:

*“Regardless if you fall into the highest category, and you tick all the boxes for all the different comorbidities, and various different underlying health issues, even if that individual says ‘I don’t want it!’, you have to honour their choice.” (6GB-S)*

*“People that work with other vulnerable people…such as healthcare workers, NHS, home care workers, you know…should give a lot of serious thought into it, shouldn’t just completely dismiss the idea…but if they’ve done their research and they know everything they think they can possibly know about it at this stage, then if they still don’t want it, then that’s their decision, it shouldn’t be compulsory.” (3GB-S)*

### (iii) Over-reach of state power

Some narratives displayed discomfort with what the coercive power displayed in any programme of mandatory vaccination potentially represented. Implicit was the question that if (UK) governments can compel vaccination, what other *abuses* of state power might that foreshadow. This was hinted at in comments which described mandatory vaccination as a *“step too far” (11GB-S), “a scary thing to think” (29GB-E)* and stated more explicitly in comments which associated mandatory vaccination with a *“huge overreach…and an abuse of power.” (20GB-W)*. Tropes which equated mandatory vaccination with totalitarian regimes (*“[like] a blooming communist country” (7GB-E)*) were deployed, particularly by those who had refused or would refuse vaccination: *“We’re free people, we’re not living in communist China, it’s not Pol Pot.” (34B-S); “You have to have a little passport to say you’ve had the vaccine before you can travel?…Yeah, it’s not what we fought the war for…That’s not freedom.” (2GB-E)*

However, participants who were not necessarily opposed to accepting COVID-19 vaccination themselves also expressed unease with the idea of states compelling others to do likewise: “*I think it will be a very, very poor society if we were forcing people to do things they didn’t wanna do” (12GB-E); “I think it’s kind of Nazi-like.” (35B-S)* It was also recognised that vaccine mandates had the potential to reinforce opposition in those already sceptical - “*it’ll just make people protest or, you know, some of the conspiracies worse” (15GB-E) -* by lending credence to the idea that management of the pandemic was motivated by a wider agenda to control the populace: *“[it will be] like, ‘oh yeah, you know, they’re, they’re trying to lock us up just for our free thinking, what is this, George Orwell’s 1984’, do you know what I mean, like, it’s just gonna fuel all of that nonsense.” (26GB-E)*

### (iv) Vaccine “apartheid”

Linked to the previous two themes were narratives concerning the potential of mandatory vaccination to create a *“two tier system” (6GB-S)*, a *“weird form of vaccine apartheid” (13GB-E)*, where the non-vaccinated were punished, barred from working in particular sectors, unable to access certain activities and services, or segregated in daily life. Again, language was often emotive, deploying metaphors which referenced extreme historical instances of segregation: “*They may as well brand us with a letter A, put a yellow star on their [sic] sleeves and say, ‘Okay, unclean, unclean.’ It’s a dreadful idea and it’ll never work” (34B-S); “We’ve been down that road through history before, and again, I think it’s very dangerous. If you take it even further, you could have people who are out on the streets…’We don’t want your family living near us, you’ve not had your vaccine.’ It could end up in all sorts of horrible situations.” (37B-S)* Such views tended to be most strongly expressed by those who were sceptical about or would refuse vaccination, particularly when imagining how their lifestyles could be curtailed if they remained unvaccinated. However, they were also voiced by some who had accepted vaccination. It was argued that there could be considerable *“detriment to society” (36B-S)* from segregating people on this basis, in the form of social unrest, vigilantism, and the potential for such division to be exploited by those in power: *R: “I think it’s a very dangerous precedent to set, and I don’t think we should go down that road at all*.

***I: When you say it’s a ‘dangerous precedent to set’, could you build me up a picture of what that would look like?***

*R: I just feel that, as a society, if that was to happen, we’d have to have a COVID passport to go into a pub or go to a shop, I just think that there is an opportunity there for someone to exploit that, to make it very divisive societally. I think it’s dangerous because you could say, ‘Well, you can’t come in because’ - I just think it gives people an excuse to divide even more and control even more, and I think that’s dangerous.” (37B-S)*

Narratives under this theme noted that anyone unable to have the vaccine because of *“underlying health issues” (47B-S)* or *“allergies” (33B-S)* should not be *“made to feel like social pariahs” (12GB-E)*. One participant noted that the costs involved in any kind of vaccine/immunity certification scheme had the potential to be discriminatory (*32B-S)*, while another stated they thought those who were less likely to be vaccinated tended to be more socio-economically disadvantaged, so any mandates could widen existing disparities, particularly if there were sanctions such as fines for non-compliance: “*I don’t think there should ever be a punitive approach [to mandatory vaccination for some occupations], particularly because a lot of the people who would be anti-vax are part of, like, lower socio-economic groups…so staff who can’t afford to pay these sort of fines. And also, fining people for things just turns you off them even more.” (26GB-E)*

While discrimination arguments were mostly used against mandatory vaccination, a few posed the counter-argument: that it was unfair for vaccinated people to suffer ongoing restrictions because others refused vaccination: *“you’re putting everybody in the same boat, that you’re not allowed to travel just because some people don’t want their vaccine, which isn’t fair.” (44B-S)*

### (v) Fluidity of support

This theme was defined by an apparent shift or variation in views in response to different scenarios or framing. Some participants moved from initial opposition to mandates to a more nuanced position which accepted that mandates may have merit in some contexts, as illustrated below. This suggests that views may not be as fixed as first appears, and that providing people with the opportunity to engage in reflection and debate reveals more nuanced responses.

*R: “I don’t think I’d like it compulsory because, again, that’s taking people’s freedom away from them…*.

***I: I think some groups have been talked about in terms of where people work, the kind of employment they have. So, if someone worked in a care home -***

*R: Oh, yes, I think that would be a good idea if you, em, for nurses, doctors and those who work in care homes. That would be a good idea. But as for in general for, you know, the population.” (24GB-E)*

*R:* “*No, I think it’s like any other vaccine, it’s the person’s choice…It’s like the flu jab, it should be like that, people can choose to get it or -*

***I: In terms of particular occupations, for example, those working with vulnerable groups, or teachers, do you think it should be a choice, or do you think it could be compulsory for certain types of jobs?***

*R: No, I think again, I still think it should be a choice…aye, I suppose you could argue for somebody who works in a care home or something else, they’re people who should be maybe vaccinated so they don’t pass on and kill all these people. That’s probably just something - a factor of where you work.” (32B-S)*

Similar internal debates were displayed in some vaccine passport narratives between, on the one hand, principles of freedom of choice, and on the other, the desire to feel safer in close proximity to other people, or, more pragmatically, simply to be able to travel again: *“I first heard about that [passports], I thought it’s wrong, but it makes sense really…if I had the injection and I went on holiday and there’s three hundred people on the plane with me and only half of them have had it, I’d…I’d be concerned…But there again I’m contradicting myself because it’s the person’s individual choice if they want that injection or not. But I would feel safer knowing that everybody on that plane was vaccinated.” (17GB-E)*

Interestingly, some participants were uncomfortable with terms such as ‘mandatory’ or ‘compulsory’, even if they agreed with the principle that vaccination should be required in some contexts. For example, one participant said she would accept hospitals saying *“If you’re going to work here, you need to have this vaccination, otherwise you can’t get employment”*, but baulked at the suggestion that staff vaccination should be *“mandatory” (31B-S)*. Another held the view *“it should be a must”* for nurses to have the vaccine but was uncomfortable with the word “compulsory”: *“it’s a wee bit of dictatorship when you start talking that way. That’s a difficult one.” (45B-S)*.

### (vi) Concerns about feasibility

Narratives under this theme focused on the operational challenges of any vaccine mandate scheme and were expressed both by those opposed to mandates and those more open to them. For the former, such concerns presented yet more arguments against mandates, while for the latter, they were issues on which they would want reassurance if any such scheme was introduced. Concerns were expressed about the cost implications (e.g. *“it’s probably going to be yet another opportunity for the government to charge an exorbitant fee” (12GB-E))*, administrative burden, potential for errors and *“mix-ups” (36B-S)*, how vaccination would be enforced and by whom *(18GB-E)*, and how complex scenarios would be handled: *“If you were going [on a flight] with a family and say there was somebody that didn’t have a vaccine passport, are they not going to be allowed to go because of their age or because of their illness? I don’t know.” (45B-S)*. It was also argued that any vaccine passport scheme would need to inspire confidence that the data held on individuals were correct - *“it’s going to be one of those things where it’s all going to boil down to how much you trust…the passport, isn’t it?” (13GB-S) -* and that passports could not be *“forged” (28GB-E), “faked” (30GB-E)* or purchased illegitimately (“*you can buy anything” (5GB-E))*.

## Discussion

This study is, to our knowledge, the first in-depth exploration of UK public views on COVID-19 vaccine mandates. It was not intended to assess levels of support for different mandates but to explore the emotions, principles and reasoning underpinning people’s views and identify contexts under which mandates may be accepted or rejected. Six themes were identified relating to (i) necessary and proportionate response; (ii) autonomy and consent; (iii) over-reach of state power; (iv) vaccine apartheid; (v) fluidity of support; and (vi) feasibility. How mandates were framed also appeared to influence response, with ‘softer’ framings such as ‘vaccination should be a requirement for employment’ evoking less unease than stark statements such as ‘vaccination should be compulsory for staff’. Because our sub-samples purposively focussed on those who, at the time of the survey (Jan-Feb 2021), indicated that they were vaccine-hesitant, we have particularly illuminated the underlying views of mandates among this important group. However, it should be noted that at the time of interview, several had accepted, or said that they would likely accept, vaccination.

Those supporting mandates or accepting them in some circumstances tended to be those who had accepted or would accept the vaccine. Unsurprisingly, those who had refused or would refuse the vaccine tended to argue strongly against mandates. However, concerns about freedom of choice, over-reach of state power and social divisiveness were not the sole preserve of the vaccine-hesitant, but were also expressed by those in favour of vaccination. The possibility that those in favour of COVID-19 vaccination might oppose its imposition on others is supported by a German study which identified around a third of respondents who were pro-vaccination but opposed to mandatory vaccination [6]. This suggests any mandate scheme which is perceived to compromise or undermine important ethical and moral principles risks jeopardising the support and trust even of those who generally support vaccination, with potential implications for future COVID-19 booster vaccination programmes.

Our study builds on research into public attitudes to mandates. Opinion surveys report mixed findings on support for various COVID-19 vaccine mandates [6,28,31]. An international survey in January 2021 found that, among 14 countries, nine supported mandatory vaccination for adults by a majority (Mexico, Brazil, South Korea, Spain, China, Italy, Canada, the UK and Australia); attitudes were divided in Japan, South Africa and the USA, while most expressed opposition in France and Germany [27]. UK poll data from December 2020 reported divided opinions on vaccine passports for air travel, leisure and public transport [25]. While in a March 2021 survey 58% supported a vaccine passport scheme while vaccine rollout was ongoing, with 34% in opposition [26]. Where public opinion is divided on an issue, it is important to understand *why* there is attitudinal diversity. Survey data provide snapshots of opinion but cannot illuminate the reasons underpinning support or opposition, or reveal through participants’ own language the emotions aroused by the issue. Qualitative research provides this insight. Qualitative views of mandates have previously been explored for childhood [32,33] and flu [34] vaccines, highlighting some similar themes (threats to autonomy, concerns about feasibility) to those in the current study. Qualitative studies of attitudes to COVID-19 vaccination have not examined mandates specifically [35–37], but provide some comparable insights, for example how acceptance can change depending on context and type of mandate [37]. Our study enabled participants to discuss and reflect in-depth, and provided detailed and timely insight into the complexity of decisions on COVID-19 vaccine mandates.

Several implications emerge from our study. The findings suggest policymakers should proceed with caution with regard to COVID-19 vaccination mandates. Even if mandates are only debated but not subsequently implemented, how the debate is conducted has the potential to arouse strong emotions and concerns. Research such as this enables policymakers to connect with public attitudes and how they may change in response to evolving circumstances. In any public debate about acceptability of mandates, context and framing are important. Some participants in our study moved from initial rejection to a more nuanced view when asked to consider the issue in more depth (for example, whether mandates could be appropriate for particular occupational groups). This suggests that engaging people in meaningful debate about specific scenarios may elicit more informed responses than simple snapshot surveys of opinion. Such methods could be deployed in public consultations to generate informed responses. It will be important to examine public attitudes towards the recent introduction of mandatory vaccination for care home workers in England and the possibility of mandates for healthcare staff [18]: for example, how concerns about staff being compelled to accept vaccination are weighed against the desire to protect vulnerable relatives in care or receiving treatment.

Reicher and Drury suggest that, if the aim of mandates is to increase vaccination uptake, the critical issue is not the response of the population in general but the potentially counterproductive effect on people who are already unsure or sceptical about vaccination [7]. An online experiment conducted before COVID-19 found that being randomised to a hypothetical compulsory vaccination intervention increased levels of anger among participants with negative attitudes to vaccination (compared with those randomised to a voluntary intervention), leading to a decrease in hypothetical vaccination uptake [38]. In our study, the highly emotive responses among vaccine refusers suggests that implementation of mandates (or even their discussion) could entrench opposition. On the other hand, some who were opposed to COVID-19 vaccination nonetheless said that the introduction of a vaccine passport scheme could persuade them, pragmatically, to accept vaccination, albeit against their wishes, in order to be able to travel abroad; something reported previously [37]. The potential longer term impact of any mandates on vaccine refusers and the vaccine hesitant (for example, whether mandates entrench or soften opposition to vaccination in future) needs to be carefully examined.

Our study has some limitations. As with all qualitative research, relatively small sample size limits the ability to draw comparisons between population sub-groups. However, our study did illuminate the factors that shaped understanding and support for mandatory vaccination, including the use of language within a key priority group, namely adults who had previously expressed hesitancy towards vaccination. Similarly, our study explored views within Great Britain and may not translate to other countries and socio-political contexts. Use of telephone interviews restricted our ability to monitor visual cues such as facial expression and body language. However, they offered participants greater anonymity (compared with face-to-face methods), which can reduce social desirability bias [39]. Such effects are likely to be more prevalent when discussing potentially sensitive topics such as vaccination behaviour and mandatory vaccination, and could explain some participants’ greater preparedness to accept vaccination mandates for occupational groups to which they did not belong. Our data were collected in two geographical regions (the whole of GB and Scotland specifically), several weeks apart. It is possible that the views of participants in the different UK nations may have been influenced by the timing of interviews and the different political and media environments. However, the same topic guide and interview sequence were used with both samples, and data were collected by the same researchers, with no evidence of differences in findings emerging between the two data sets. Finally, we did not explore views of mandatory COVID-19 vaccination for children [40,41], an issue which continues to be debated publicly [42].

## Conclusions

Mandating any vaccination involves consideration of complex issues and provokes strong views. Our findings illustrate the potential to entrench strong views further and increase vaccine hesitancy. Debate about mandating COVID vaccine should be informed by detailed investigation of public views and approached cautiously.

## Supporting information

SRQR Checklist

CoI declaration

CoI declaration

CoI declaration

CoI declaration

CoI declaration

CoI declaration

CoI declaration

CoI declaration

CoI declaration

CoI declaration

CoI declaration

CoI declaration

## Data Availability

The anonymised survey data are available upon reasonable request to the authors

## Author contributions

Conceptualisation, MS, AF, DE, AMM, AM, CJ, HB, KH; data collection and analysis, MS, AF, DE, HB, CE; writing – original draft MS AF; writing – review and edition, MS, AF, DE, HB, AMM, AM, HB, KH, MU, KA.

## Ethical approval

Stirling University General University Ethics Panel (GUEP 2021 1002) provided approval for all elements of the qualitative study. The OPTIMUM survey received approval from NatCen’s Research Ethics Committee (ID P14307). All participants gave informed consent before taking part.

## Acknowledgements

We thank all the participants who volunteered for the study and shared their experiences and views, and Irene Miller at ScotCen who assisted with fieldwork.

